# Policies, practices, and experiences of European biobanks on sharing genomic biobank results with donors - a survey of BBMRI-ERIC biobanks

**DOI:** 10.1101/2025.09.25.25336629

**Authors:** Minna Brunfeldt, Terry Vrijenhoek, Helena Kääriäinen

## Abstract

To study European biobanks’ policies, practices, and experiences on communicating individual research results to participants the EU Horizon 2020 Project ‘Genetics Clinic of the Future’ performed two surveys in 2016 and 2020. First, a questionnaire was sent in 2016 (Survey I) to 351 European biobanks in 13 countries that were members of Biobanking and Biomolecular Resources Research Infrastructure – European Research Infrastructure Consortium (BBMRI-ERIC). We received replies from 72 biobanks (response rate 21%), representing each of the 13 BBMRI Member States. Respondents were mainly directors or heads of biobanks. To evaluate how the policies and practices of biobanks evolved over time, we also conducted another survey in 2020 (Survey II). The Survey I was implemented using a web based Webropol tool, and the Survey II was distributed by email. The biobanks had very different policies of sharing genomic data and the policies had changed over time. The percentage of biobanks with a policy to share results with participants if they so wish had increased between 2016–2020 from 36% to 45%. On the contrary, the percentage of biobanks with a policy to pro-actively re-contact the participants to share (some) results had decreased from 52% to 39%. Still in 2020, half of the biobanks had never shared results with participants.

## Introduction

Research findings including genomic data of the participants are growingly accumulating in biobanks as the new technologies have strongly promoted use of various types of genome wide analysis in biobank research^1^. The discussion about whether this data should be used, in addition to research, for promoting health of the individual donors started two decades ago and is continuing^2–4^. In spite of the several examples of the health benefits that personal genomic data can bring to the individuals, the discussion around sharing biobank results with donors presents contradictory opinions^5^. Several problems have been recognized. If biobanks started increasingly sharing genomic results with participants, they would need resources to do that^6^. In addition to the work relating to the mere reporting, a thorough investigation of several issues would have to be carried out. Can the results from biobank studies be considered reliable? Which type of results could the biobank return? How the information and, when needed, genetic counselling of the donors could be organized? These and other questions have been discussed and no uniform answers have emerged^7,8^. In addition, sharing results might put pressure on healthcare, since some of the participants may need medication or other treatment^9^. For example, in the study of Marjonen et al. 2021, where 3,177 biobank participants received feedback on their individual genetic risk of coronary heart disease, type 2 diabetes and venous thrombosis, the individuals that got high risk scores for any of the conditions were advised to contact their healthcare provider, aware of the fact that it might cause burden for the healthcare^10^. In spite of the doubts and problems listed above, there also are data and experience to support sharing biobank results. According to the systematic review of stakeholder opinions^11^ there is overwhelming evidence of high interest in return of results from potential and actual genomic research participants and there are studies suggesting that sharing results seems not to cause adverse effects^12^. However, health professionals and especially genetics professionals have had more conservative views compared with other stakeholders^13^. An argument for sharing results might also be that some screening programs based on polygenic risk scores seem to be cost-effective. For instance, the article by Wong et al. 2021^14^ describes that a breast cancer screening programme that incorporated genetic testing was more cost-effective than a regular mammogram-only screening programme^15^. From our previous study we know that the legislation concerning the sharing of biobank results with participants in different EU countries is either varying or there is no legislation related to this^16^.

Here we report the results of the investigation performed by the EU Horizon 2020 Project ‘Genetics Clinic of the Future’ to learn about the policies, practices and experiences of European biobanks in two different situations: actively offering individual research results from biobanks to donors or responding to the possible requests from donors for their results. The goal was to gain better understanding of the possible similarities as well as differences between European biobanks^16^. This is important especially as European biobanks have several large collaborations and it might be confusing and even unfair if donors participating in the same collaborative project are treated differently in this respect.

## Materials and Methods

Informal in-depth discussions among the interdisciplinary GCOF consortium (at two F2F Project Meetings and subsequently via email) formed the basis for preparation of this questionnaire survey. All biobanks listed at that time on the BBMRI-ERIC Directory 1.0^17^ representing each of the 13 BBMRI Member States were invited to join the Survey and the responses were collected in the beginning of 2016. The questionnaire was conducted in English for all the respondents. The questionnaire was formulated using the Webropol online survey and analysis tool^18^. The BBMRI-ERIC network advertised the survey through their web page and Newsletter. In addition to multiple choice questions, also all free comments were collected and considered. In 2020, we implemented a brief second survey (Survey II) four years after the first survey to understand longer-term developments in the policies and practices of European biobanks on sharing genomic results with participants. The email questionnaire was sent to all the biobanks which had responded to the Survey I. When we analyzed the change between Survey I (2016) and Survey II (2020), we compared the replies from only those biobanks which had responded to both Surveys.

## Results

In the BBMRI-ERIC Directory 1.0, there were 511 contact addresses for biobanks. After the removal of duplicated addresses, the questionnaire was sent to 380 biobanks in 13 countries, of which 24 addresses did not work and five biobanks replied that they did not have genomic data, human samples or that they were only small cohorts. This left 351 biobanks in total. Seventy-two replies were received (response rate 21%) to Survey I, which represented all 13 BBMRI-ERIC countries at that time (Table 1). Of those 72 biobanks, we reached 33 in 2020 for the Survey II. The responding biobanks represented a variety of different human collections; the respondents categorized their biobanks as one or more of the following: 45/72 (63%) clinical biobanks, 36/72 (50%) disease-specific collection, 24/72 (33%) population cohorts, 7/72 (10%) other type of collection and 5/72 (7%) collection derived from clinical trials. Most of the respondents, 66/72 (92%) respondents identified themselves as biobank directors, heads, managers or similar.

**Table 1.** The number of the respondents by country.

| *Member State* | *Respondents*<br>*2016* | *reply-%* | *Respondents*<br>*2020* |
| --- | --- | --- | --- |
| *Czech Republic* | 4/5 | 80 | 0/4 |
| *Italy* | 18/64 | 28 | 7/18 |
| *France* | 10/70 | 14 | 3/10 |
| *Malta* | 1/1 | 100 | 0/1 |
| *Belgium* | 10/14 | 71 | 5/10 |
| *Austria* | 1/3 | 33 | 1/1 |
| *Netherlands* | 9/107 | 7 | 5/9 |
| *Finland* | 3/8 | 38 | 3/3 |
| *Poland* | 1/2 | 50 | 0/1 |
| *Germany* | 5/10 | 50 | 2/5 |
| *Estonia* | 1/1 | 100 | 1/1 |
| *Greece* | 1/1 | 100 | 0/1 |
| *Sweden* | 8/65 | 12 | 6/8 |
| *Together* | **72/351** | **21** | **33/72** |

When asked in Survey I about the policies concerning actively contacting the participant (biobank’s initiative on sharing results), more than half 39/72 (54%) of the biobanks informed that they did not have a policy whether or not to proactively share the results with participants. On the other hand, 27/72 (38%) of the biobanks had a policy to proactively share the results with participants. A few biobanks 6/72 (8%) had a policy not to share results proactively with participants (Figure 1). Additionally, about half of the biobanks 37/72 (51%) replied that they had a policy to return (some) results to participants if they so request. A few biobanks 10/72 (14%) had a policy not to share results even if requested by the participants. Thus biobanks appeared to be more willing to share the results if the participants requested when compared with their willingness to proactively contact the participants for offering to share results (Figure 1).

**Figure 1.**
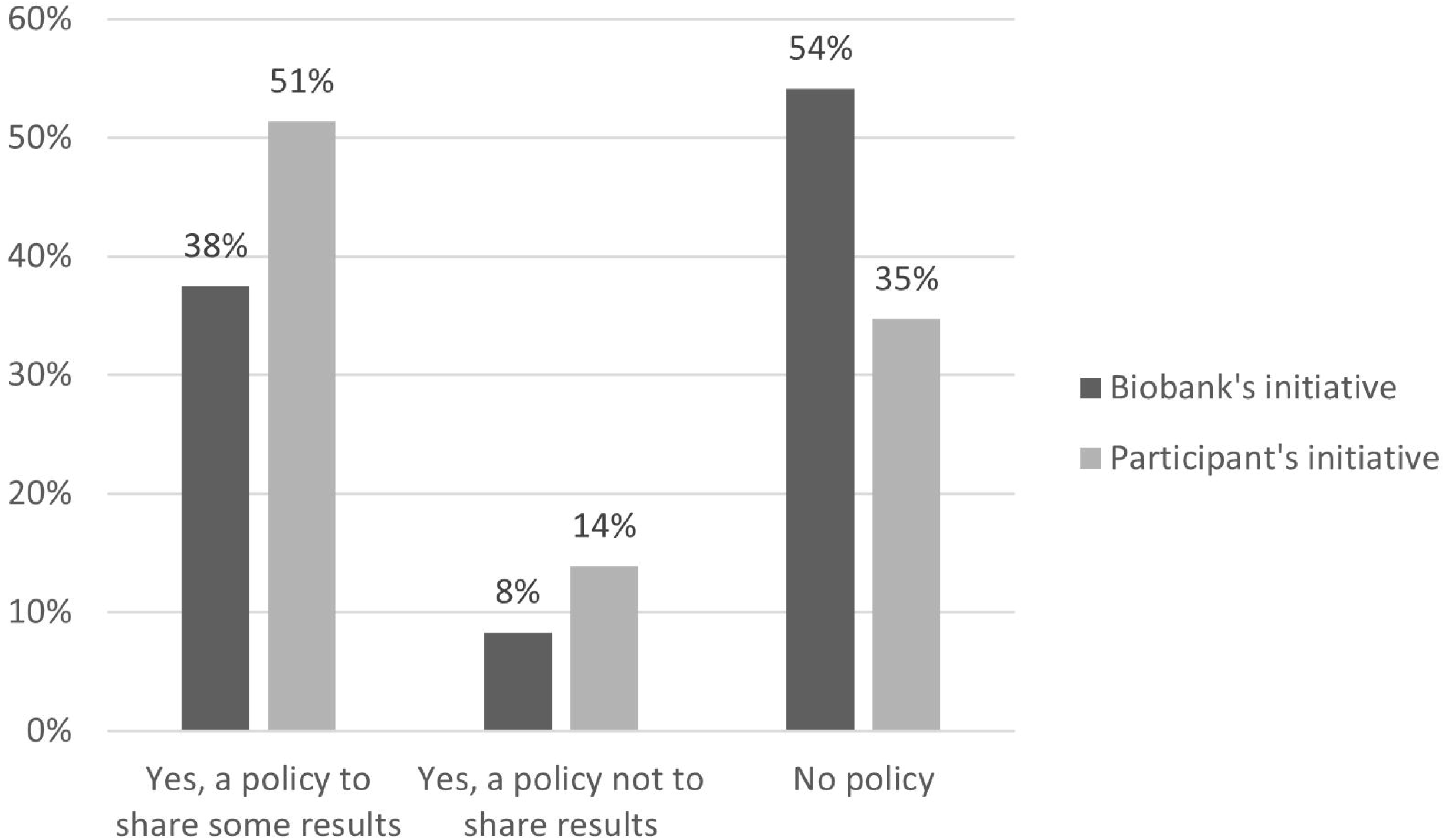
Replies to Survey I (2016) question (Biobank’s initiative): “Does your biobank have a policy about pro-actively re-contacting participants to share (some) results that might benefit them?” and question (Participant’s initiative): “Does your biobank have a policy about sharing results with participants if they so request?” (n=72).

Of the 37 biobanks that had a policy to share results with participants when they so request, 12/37 (32%) would share any specific results requested with no limitations, 13/37 (35%) would share requested results only if the biobank considered them beneficial to the health of the participant. Of the biobanks 3/37 (8%), would give all existing genomic data when requested.

Of the 27/72 (38%) biobanks with a policy to pro-actively share results to participants, 6/27 (22%) would share results related to potentially fatal, but treatable monogenic diseases or disease risks and 7/27 (26%) would share results related to family planning.

Over half of the biobanks 17/27 (63%) mentioned that the results would be validated in a clinical laboratory before returning them to the participant.

The 27 biobanks which would pro-actively share results with participants responded that the recommended ways to contact participants to inform results were by phone 18/27 (67%), by letter 13/27, (48%) and by email 11/27 (41%).

For more information about the consequences of the results or possible treatments, most of the biobanks would refer the participants to a public clinic instead of a private one, regardless of whether it was the participant’s initiative to request results or biobank’s initiative to report results. For instance, in situations where participants are requesting results, 14/37 (38%) of the biobanks that had a policy to share results with participants when they so request would refer the participant to a public genetic clinic for further information, and 15/37 (41%) to a public specialist clinic relating to the result, while only 4/37 (11%) would refer the participant to a private genetics clinic and 3/37 (8%) to a private specialist clinic relating to the result (Figure 2).

**Figure 2.**
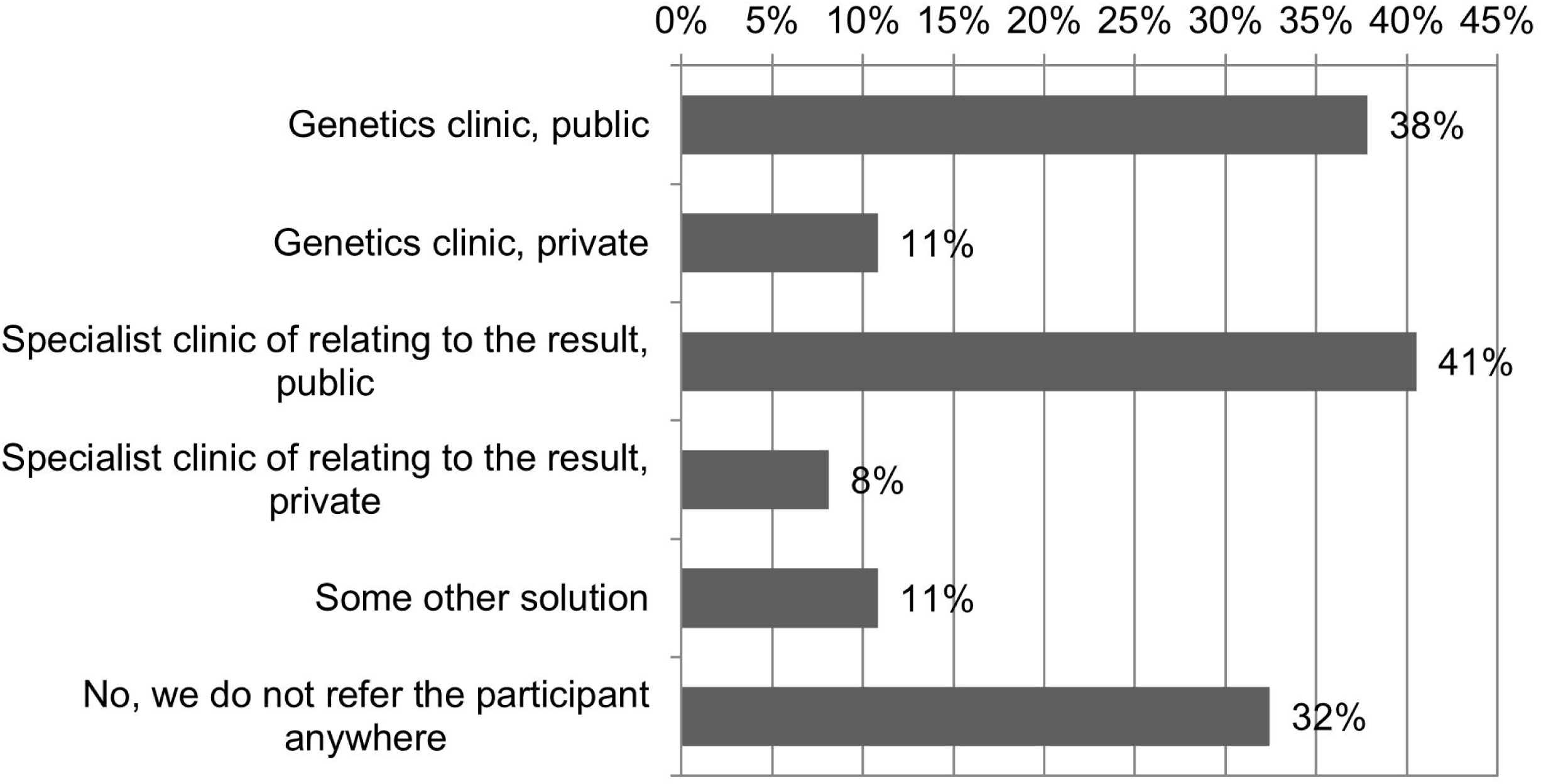
Replies to Survey I (2016) question: (Participant’s initiative) “Are participants referred for more information or possible treatment relating to the biobank results?” (n=37). The respondents were allowed to choose several options.

In between Survey I (2016) and Survey II (2020), more biobanks had defined their policies so that the number of biobanks without a policy, had decreased over time (Figures 3 and 4). The amount of biobanks, with a policy not to share the results with participants, had notably increased. In line with that, fewer biobanks had a policy to proactively re-contact participants to share results and more biobanks had a policy not to proactively re-contact participants to share results (Figure 3). By contrast, there was a polarization of the policies regarding sharing of the results when requested: fewer biobanks had no policy, and there were both more biobanks that had a policy to share results when requested and more biobanks that had a policy not to share results when requested (Figure 4).

**Figure 3.**
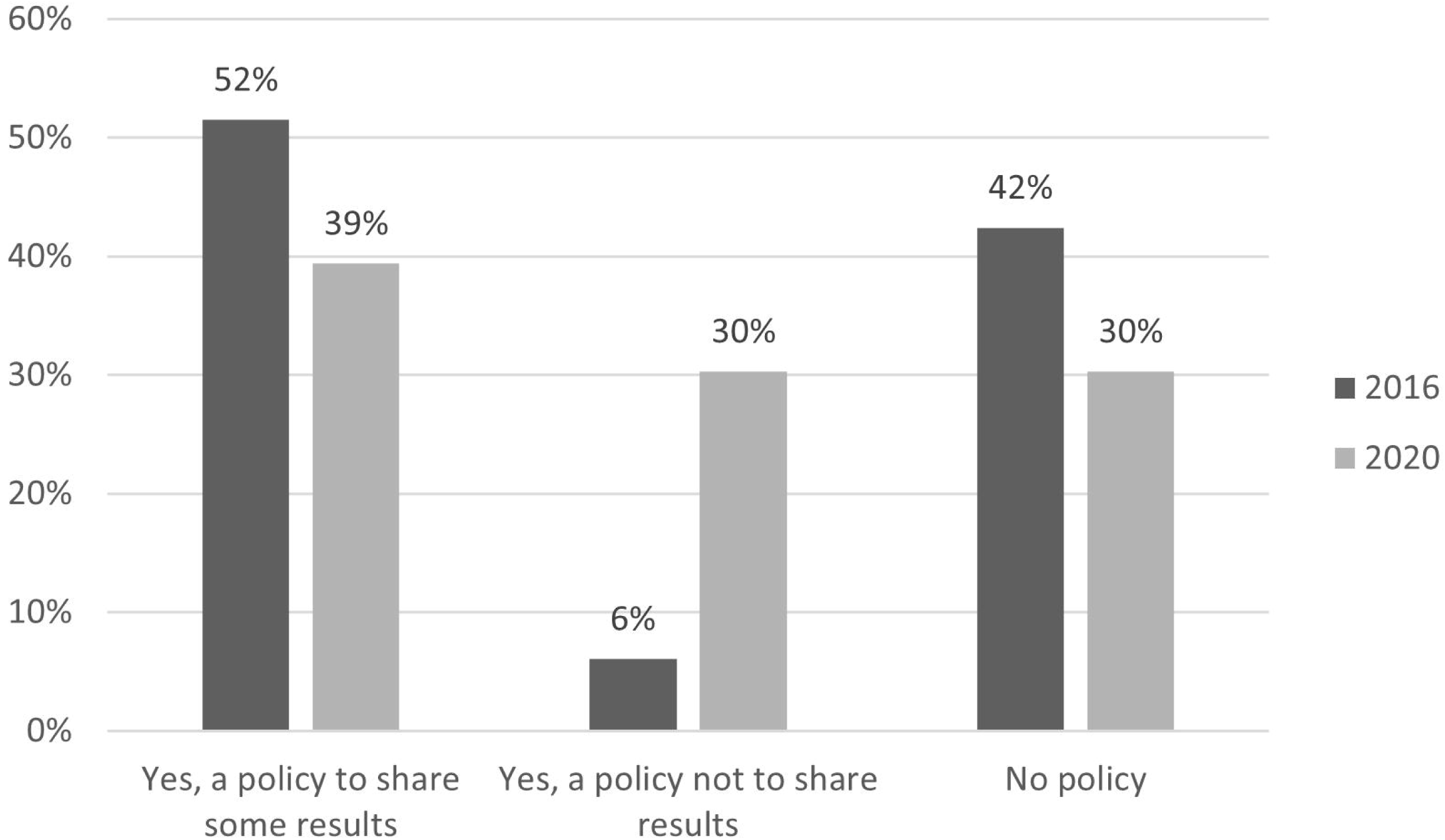
Biobank’s initiative, Survey I and Survey II. Replies to question: “Does your biobank have a policy about pro-actively re-contacting participants to share (some) results that might benefit them?” n=33.

**Figure 4.**
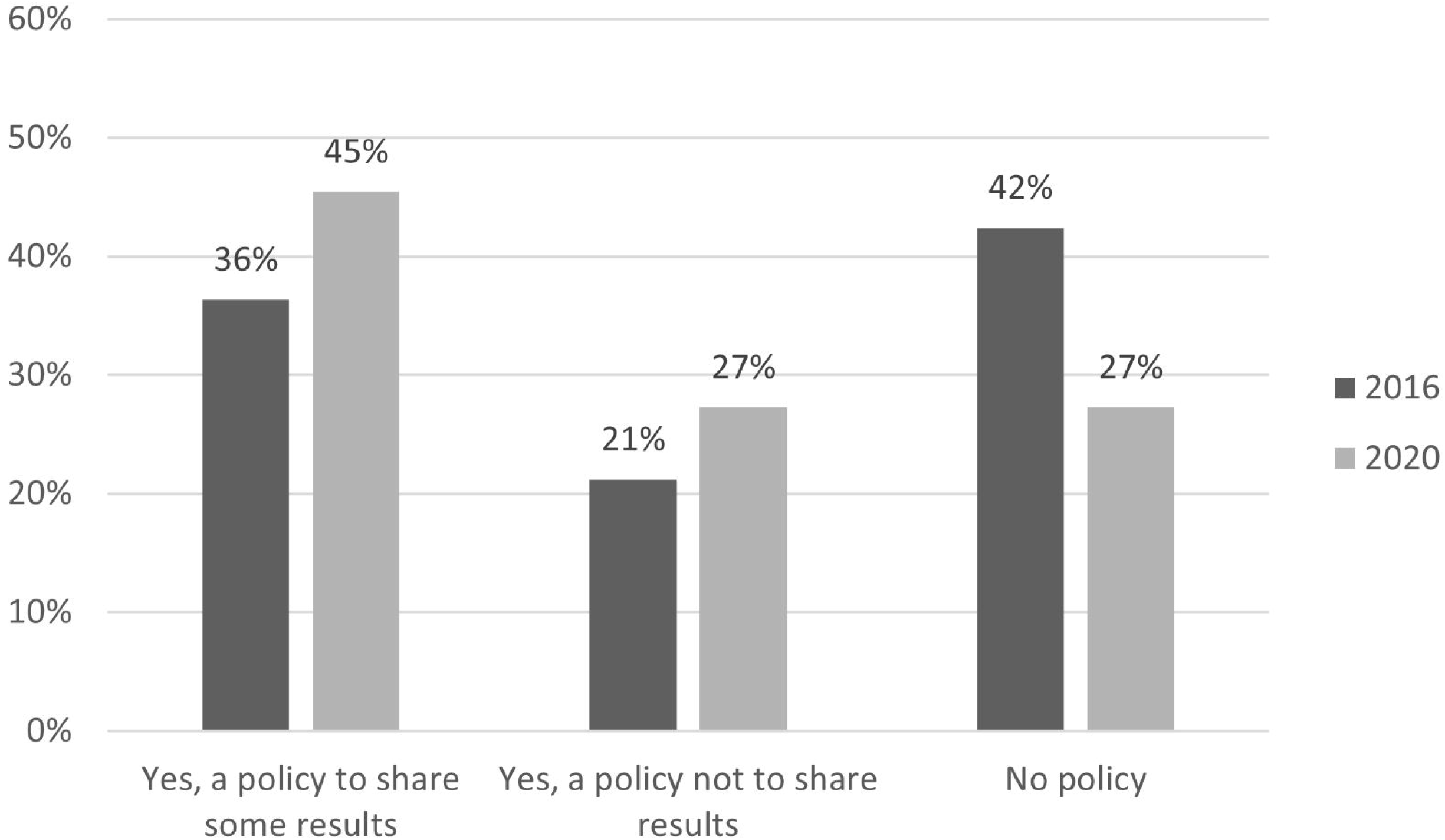
Participant initiative Survey I and Survey II. Replies to question: “Does your biobank have a policy about sharing results with participants if they so request?” n =33.

Of the biobanks that replied in Survey I (2016) to have a policy not to pro-actively inform results to participants 6/72 (8%) and/or to have a policy not to share results with participants when they so request 10/72 (14%), the most frequent explanation for this was that they had difficulties with assessing the health benefit 6/10 (60%).

At the time of the Survey II (2020) biobanks replied that they had shared results with participants more frequently than earlier (Figure 5). It seemed that especially the number of biobanks sharing results to participants when they so request had increased. The majority of the biobanks had never shared results with the donors, proactively by the biobank 22/33 (67%) nor when the participants request 21/33 (64%). These biobanks represent those with policies to share as well as those with policies not to share and those with no policies.

**Figure 5.**
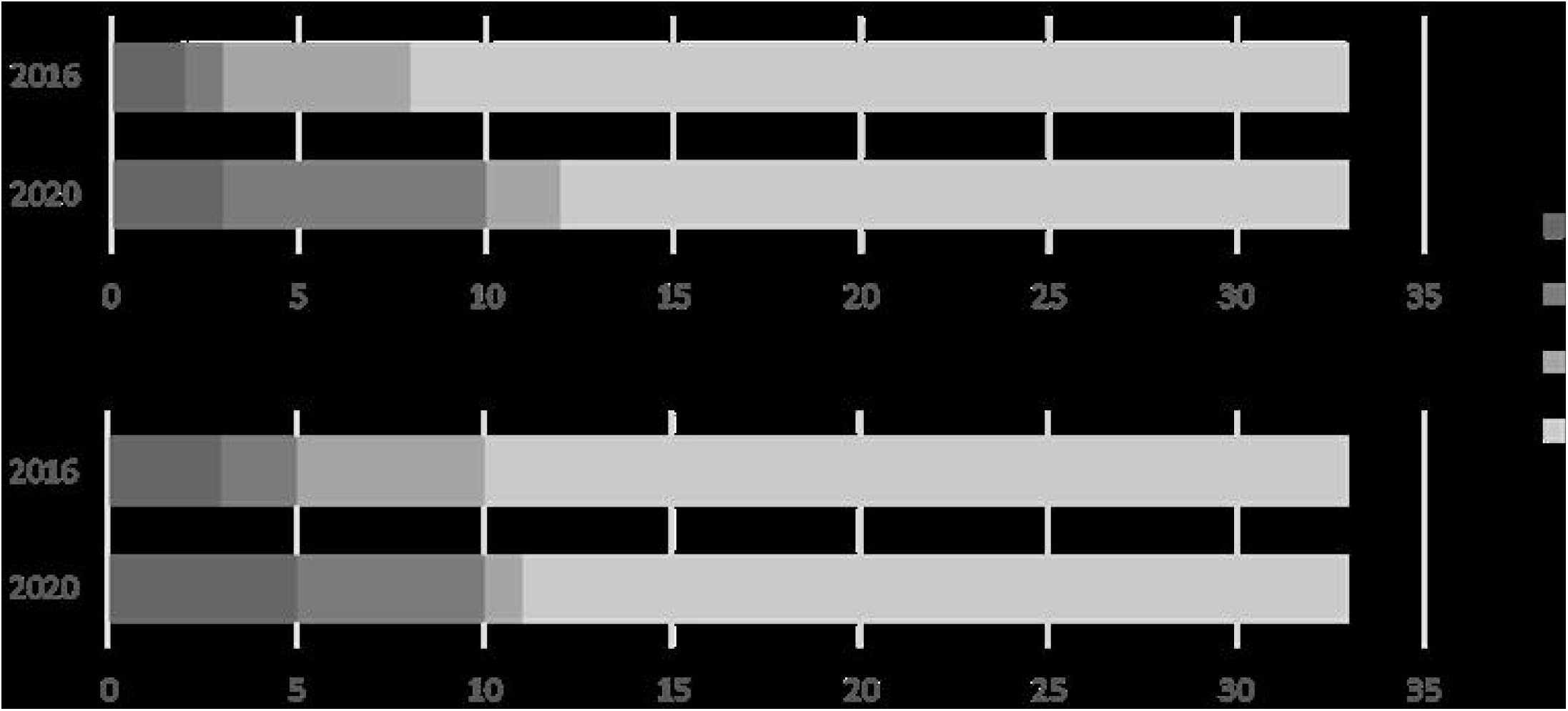
Replies to questions; Participant’s initiative, “How often have you shared biobank results with participants?” and Biobank’s initiative, “How often have you shared biobank results with participants?” n=33.

## Discussion

To investigate European biobanks’ policies, practices and experiences on communicating individual research results to participants and the possible change in them the EU Horizon 2020 Project ‘Genetics Clinic of the Future’ performed two surveys among BBMRI biobanks, one in 2016 and the other in 2020. The response rate to Survey 1 was 21%. One of the reasons for the rather low response rate probably was the heterogeneity of the BBMRI biobanks ranging from population collections and birth cohorts to hospital samples, case-control studies and even non-human collections. This diversity of biobanks may have been especially big in countries like the Netherlands and Sweden with a very high number of biobanks listed in the BBMRI Directory and a very low response rate (9/107 and 8/65, respectively). According to the Directory, it could be assumed that not all listed biobanks had DNA nor genomic results. Thus many BBMRI biobanks maybe immediately felt that the Survey did not concern them. Survey 2 was sent only to those biobanks that had replied to Survey 1. We do not know why about half (54%) of these biobanks did not reply to Survey 2.

There are several examples of pilot studies offering genomic results from biobanks to the donors. In the P5 study^10^ the possibility to get genomic results was offered to 8217 individuals, participants of the population based FinHealth 2017 study, and 3177 (56%) accepted the invitation. This indicates that the interest in shared genomic results is high, at least in this Finnish population. In line with this, our earlier study by Brunfeldt et al. 2022 found that about half of the patients and family members from an international diagnostic laboratory Blueprint Genetics opted in to receive actionable genomic results as part of diagnostic whole-exome sequencing^19^. There have been several recent pilot studies evaluating the possible benefit of using genomic data, including polygenic risk scores, for the health promotion of biobank donors. Also, the usability of new methods for sharing results with large numbers of research donors has been piloted^10,20,21^.

In our earlier study, 72% of the European biobanks told that their national legislation would allow them to contact their participants in order to inform them about their results^16^. Over half of the European biobanks (56%) also reported that they mention in their consent process that the participants have a possibility to request their results from the biobank (3). With this background, according to the present study surprisingly few biobanks had policies for these possibilities: only 38% of the biobanks had a policy to proactively share the results and only 51% had a policy to share if the participant so requested.

Our follow up study suggests that the European biobanks have growingly (Figure 4) formulated policies to share results when the request comes from the participant. In addition to possible health benefits, this is also expected to better engage the donors to the biobank and to research in general^20^. On the contrary, the proportion of the biobanks with a policy to proactively share results with participants seems to be decreasing (Figure 3). This is understandable as the decision to share results is a complex one. The utility of actively sharing genetic or genomic results includes several components including the analytical and clinical validity of the result, possible health outcome, personal utility, equity, feasibility and possible economic impact^22,23^. But surprisingly, 35% of the biobanks we surveyed had no policy whether to share the results to study participants upon request. This means that if their participants requested for their results, they would not have a predetermined way of answering their requests. Similarly, they appeared not to have a solution to a situation where the biobank would consider reporting of some findings beneficial for their donors’ health (Figure 1). When comparing our European data with a recent study about the policies of biobanks in

United States interesting differences emerge^24^. Of the 327 biobanks in the U.S. survey, 62% of biobanks had a policy about sharing results. Of the policies, 57% stated that results would never be returned. Surprisingly, while European professional opinion seems to be more restrictive than that in U.S.^5^, the U.S. biobanks appear to be more cautious about returning results than European biobanks.

Figure 5 shows how often the biobanks had actually shared results. It seems that the participants did not use their right to know their results very actively. In addition, it appears that biobanks proactively shared results less actively than they potentially could, based on their policies. Goisauf et el. 2019 surveyed biobank professionals and revealed a need for practical, tangible and hands-on ethical and legal guidance for possible sharing of results^25^. In the Editorial of a resent Special Issue: “Genetic Counseling and Genetic Testing in Precision Medicine”^26^, the need for new approaches to accessing genetic counseling and genetic testing in addition to the genetic counselling performed in specialized tertiary healthcare settings was strongly emphasized. According to Survey 1, one of the practical obstacles is the need for the validation of every result from a new sample in a clinical laboratory. This is considered to be difficult, slow and expensive. Also, biobanks might not have enough resources for contacts and information needed for sharing of results. Electronic communication like the web portal for communicating polygenic risk scores^10^ or integrating a genetic report into electronic health records^21^ are solutions worth considering and developing further. Traditionally, these genetic services were only available in specialized tertiary healthcare settings for those with suspected genetic conditions or a family history of such conditions. However, new approaches to accessing genetic counseling and genetic testing are being investigated.

In addition to biobanks and research projects, individuals may get genomic results from direct-to-consumer (DTC) companies. This could aggravate health inequality, as not all consumers are able to afford DTC genetic testing^7^. The risk of inequality might also be the reason why many biobanks responding to Survey I mentioned referring biobank participants, after having received genomic results, to a public clinic instead of a possible private one for further information. There might be a lack of private option in some of the BBMRI countries.

In spite of the expected boost to health promoting that genomic results from biobanks might offer, returning individual results to biobank donors was still very rare. Summarizing the lessons learned from pilot studies might reveal the gaps in the knowledge that still have to be filled in with further studies. Europe with its high-quality public health care offers an optimal field for such studies.

### Strengths

To our knowledge, this is the most comprehensive survey to date of the policies of European biobanks to share genomic results with their participants, which provides a broad view on the diversity of such policies.

### Limitations

Not all biobanks replied to Survey I, which limits the generalizability of the findings of our study. Further, of the 72 biobanks that replied to the Survey I in 2016 only about half (46%) participated in the Survey II in 2020, which limits our ability to examine the evolution of biobank’s result sharing policies over time. Another limitation is that while we received several replies from some countries (i.e. Italy), only very few replies were received from some other countries (i.e. the Netherlands). This disparity partly stems from the difference in biobanking structure in different countries.

## Conclusions

The policies of European biobanks about sharing genomic results with participants differ widely from proactive policies to share results to policies not to share results even when requested and to having no policies at all about the matter. Half of the biobanks had never shared results with their participants. This indicates that the possible health benefits that the returning of results might offer do not materialize.

## Data Availability

The data of this study are not publicly available as participants did not provide consent for their data to be shared.

## Acknowledgements

We thank BBMRI-ERIC for distributing our Survey to the BBMRI-ERIC Members in their channels. Also, we kindly thank all the biobanks that participated in the Survey. We warmly thank all the members of the GCOF project.

## Credit Author Statement

**Minna Brunfeldt:** Conceptualization, Writing- Original draft preparation. **Terry Vrijenhoek:** Writing- Reviewing and Editing. **Helena Kääriäinen:** Conceptualization, Writing- Original draft preparation, Supervision.

## Ethics Review Statement

Ethics committee approval was not required for this questionnaire study.

